# Vasopressor Modulation of Cerebral Autoregulation and Systemic Hemodynamics in Severe Traumatic Brain Injury: The REGULATE randomized controlled trial

**DOI:** 10.1101/2025.10.02.25337200

**Authors:** Athanasios Chalkias

## Abstract

**Background:** Severe traumatic brain injury (TBI) frequently disrupts cerebral autoregulation, rendering cerebral perfusion pressure highly dependent on systemic hemodynamics. Vasopressors are a cornerstone of TBI management, yet norepinephrine and vasopressin may exert divergent effects on cerebral autoregulation and systemic cardiovascular function. Their comparative impact in this population remains poorly defined.

**Objective:** The REGULATE trial is a randomized, controlled study designed to investigate the effects of norepinephrine, vasopressin, and their combination on cerebral autoregulation and advanced systemic hemodynamics in patients with severe TBI.

**Methods:** This quadruple-blinded, parallel-group trial will enroll 450 adults with severe TBI (Glasgow Coma Scale ≤ 8) requiring vasopressor support. Participants will be randomized (1:1:1) to norepinephrine, vasopressin, or combination therapy. Continuous multimodal monitoring will include intracranial pressure, arterial pressure, near-infrared spectroscopy, and advanced cardiac output assessment. Co-primary outcomes are (1) cerebral autoregulation, quantified as the area under the curve of the pressure reactivity index over 48 hours, and (2) systemic hemodynamics, defined as the area under the curve of cardiac output and effective arterial elastance over 48 hours. A prespecified interaction analysis will evaluate treatment effects on the relationship between cerebral autoregulation and systemic hemodynamics. Secondary outcomes include cerebral oxygenation indices, intracranial pressure burden, vasopressor dose exposure, organ perfusion markers, adverse events, and clinical outcomes (mortality, ICU length of stay, neurological recovery at 3 months).

**Conclusions:** REGULATE is the first adequately powered trial to systematically compare norepinephrine, vasopressin, and their combination on cerebral autoregulation and systemic cardiovascular performance in severe TBI. Results are expected to inform individualized vasopressor strategies to optimize cerebral and systemic physiology while minimizing secondary injury.

**Trial registration:** ClinicalTrials.gov (forthcoming).

## 1. BACKGROUND

Traumatic brain injury (TBI) often leads to significant disruption of cerebral autoregulation – the intrinsic ability of cerebral blood vessels to maintain stable cerebral blood flow (CBF) despite fluctuations in systemic blood pressure. Impaired autoregulation contributes to secondary brain injury by rendering cerebral perfusion pressure (CPP) more dependent on systemic hemodynamics, increasing the risk of both ischemia and hyperemia [1,2]. The degree of autoregulatory impairment has been associated with worse outcomes, including higher mortality and poor neurological recovery [3,4]. Monitoring tools such as the pressure reactivity index (PRx) and other dynamic cerebrovascular indices have been developed to assess autoregulation at the bedside, enabling individualized CPP targeting in some centers [5,6]. But for these patients, controlling CPP and mean arterial pressure (MAP) using vasopressors continues to be one of the most crucial therapeutic interventions.

Norepinephrine and vasopressin, both used to raise blood pressure, have distinct effects on cerebral autoregulation. Norepinephrine primarily constricts blood vessels, potentially impacting cerebral autoregulation by reducing blood flow to the brain at higher doses or in certain conditions [7]. While norepinephrine can increase MAP and improve cerebral blood flow velocity, it can also impair cerebral autoregulation at higher doses or in specific situations. Studies have shown that in healthy volunteers, norepinephrine increases pulsatility index but has no effect on cerebral autoregulation or cerebrovascular reactivity to carbon dioxide [7-12]. On the other hand, in cases of TBI, excessive norepinephrine-induced vasoconstriction could exacerbate ischemic damage by reducing blood flow to already vulnerable brain tissue [7,13-16].

Vasopressin primarily acts on V1 receptors, causing vasoconstriction, and may theoretically have a more targeted effect on cerebral vessels. However, its effects on different vascular beds, including the brain, vary [17-20]. The impact of vasopressin on cerebral autoregulation is less clear compared to norepinephrine. Some studies suggest it may be less likely to impair autoregulation than norepinephrine [10,11,13,18,21]. Nevertheless, the impact of vasopressin on cerebral autoregulation can vary depending on the clinical context, such as the presence of brain injury, sepsis, or other comorbidities [11,13], with higher doses potentially impairing autoregulation [7,11].

Despite achieving similar hemodynamic parameters and CPP, the effects of norepinephrine and vasopressin on cerebral oxygenation, CBF, and metabolism are heterogeneous and not fully understood [13,22]. In addition, therapeutic strategies to support or restore autoregulatory function remain limited [23-25]. In this context, understanding how norepinephrine and vasopressin influence cerebral autoregulation is essential for optimizing hemodynamic management and minimizing secondary injury.

## 2. AIM

The REGULATE is a quadruple blind, parallel group, open-label randomized controlled trial aiming to investigate the effects of norepinephrine, vasopressin, and their combination on cerebral autoregulation and systemic hemodynamics in patients with severe TBI.

## 3. METHODS

### 3.1 Study design

Patients with severe TBI who are admitted to the Intensive Care Unit (ICU) will be included. Ethical approval will be provided by the Ethics Committee of the Tzaneio General Hospital. The study will be designed in accordance with the declaration of Helsinki and will be registered at ClinicalTrials.gov. An independent steering committee and independent data and safety monitoring committee will review the trial regularly, assessing conduct, progress, and safety. Written informed consent will be obtained from all patients or next-of-kin in case of death or if the patient remains incompetent to give consent.

### 3.2 Patient eligibility

Patients fulfilling the following criteria will be included: (a) Age ≥ 18 years;(b) Diagnosis of severe TBI, defined as Glasgow Coma Scale (GCS) ≤ 8 after resuscitation; (c) Admission to Emergency Department (ED) or ICU within 12 hours of injury; (d) Clinical indication for vasopressor support to maintain CPP or MAP per institutional protocol; (e) Invasive monitoring available and feasible, including intracranial pressure (ICP) monitoring and arterial line for advanced hemodynamics; (f) Written informed consent obtained from patient’s legally authorized representative.

Exclusion criteria will be: (a) Known pregnancy; (b) Pre-existing severe cardiac disease (e.g., severe heart failure, cardiogenic shock, significant valvular disease) that would preclude study vasopressor use; (c) Severe systemic vasculopathy (e.g., critical limb ischemia, severe peripheral artery disease) limiting vasopressor safety; (d) Known allergy or contraindication to norepinephrine or vasopressin; (e) Non-survivable TBI or expected death within 24 hours; (f) Active participation in another interventional clinical trial that conflicts with vasopressor administration or cerebral monitoring; (g) Intracranial pathology unrelated to TBI likely to independently affect cerebral autoregulation (e.g., recent stroke, intracerebral tumor, or arteriovenous malformation); (h) Severe chronic kidney or liver failure (e.g., CKD stage 4–5, Child-Pugh C) that could alter vasopressor metabolism or outcomes.

The inclusion and exclusion criteria are designed to select a homogeneous population with severe TBI requiring vasopressors, ensure the safety and feasibility of advanced hemodynamic and cerebral monitoring, and exclude conditions that could confound autoregulation or hemodynamic responses.

### 3.3 Randomization and blinding

All patients will be screened for eligibility at the time of vasopressor commencement (ED or ICU). All clinical staff in both clinical areas will be provided with trial education for the duration of the trial, and will be empowered to screen and enroll eligible patients.

A computer-generated permuted block randomization list with varying block sizes will be concealed and created by a statistician. Eligible patients will be randomized in a 1:1:1 ratio to 0.1-0.5 μg kg^-1^ min^-1^ norepinephrine infusion (Group N), 0.01-0.06 IU m^-1^ arginine vasopressin (AVP) infusion (Group V), or norepinephrine (0.1-0.5 μg kg^-1^ min^-1^) plus AVP (0.01-0.06 IU m^-1^) infusion (Group NV). A vasopressor infusion will be defined as the continuous administration of a vasoconstrictor agent (any one of noradrenaline, vasopressin, or their combination) via an infusion pump.

Given the nature of the intervention, blinding of treating clinicians is not feasible; therefore, the trial will be conducted in an open-label fashion. However, a quadruple blinding will be followed, i.e., the treatment assignment will be concealed from subjects/families, outcome assessors, data analysts, and staff members performing the follow-up.

### 3.4 Procedures

All included patients will receive standard medical care as determined by the treating clinicians and according to international guidelines [26-30]. Intracranial pressure will be monitored and the amount and type of fluids and the dose of vasopressors will be titrated to maintain CPP between 60-70 mmHg [31,32]. Therapeutic intensity level and the Sequential Organ Failure Modified Oxford Handicap Scale (MOHS) will be assessed as previously described [33-35]. Mortality from any cause will be included.

#### 3.4.1 Systemic hemodynamics

All patients will be managed using standard, evidence-based practices according to recent guidelines.

The internal jugular or subclavian vein will be cannulated with a triple-lumen central venous catheter that will be connected to another pressure transducer to measure central venous pressure (CVP) and central venous oxygen saturation (ScvO_2_). The radial artery will be cannulated and connected to an advanced clinical platform to directly measure systemic hemodynamic parameters including systolic (SAP), diastolic (DAP), and MAP, CO and cardiac index (CI), stroke volume (SV), stroke volume variation (SVV), and systemic vascular resistance (SVR). Before making each measurement, we will confirm that transducers are correctly leveled and zeroed [36,37], while the system’s dynamic response will be confirmed with fast-flush tests [38]. Damping will be assessed via repeatedly performed visual inspections of the pressure waveform at the end of a fast flush test. Artifacts will be detected and removed when documented, and when measurements are out-of-range or SAP and DAP are similar or abruptly changed (≥40 mmHg decrease or increase within two minutes before and after measurement).

Critical care transthoracic echocardiography will be performed twice daily by a highly experienced echocardiographer who will be blinded to the individual and study sequence. Left ventricular ejection fraction (LEVF), left ventricular outflow tract (LVOT), and LVOT velocity time integral (LVOT-VTI) will be recorded among others. All echocardiographic parameters will be calculated from five measurements (regardless of the respiratory cycle) and analyzed retrospectively.

#### 3.4.2 Determinants of venous return

The methods of the mean circulatory filling pressure (Pmcf) analogue (Pmca) and related values algorithm have been described in detail before [39-42]. Briefly, based on a Guytonian model of the systemic circulation [CO = *V*R = (Pmcf − CVP) / RVR], an analogue of Pmcf can be derived using the mathematical model Pmca = (a × CVP) + (b × MAP) + (c × CO). In this formula, a and b are dimensionless constants (a + b = 1). Assuming a veno-arterial compliance ratio of 24:1, ‘a’ = 0.96 and ‘b’ = 0.04, reflecting the contribution of venous and arterial compartments, and ‘c’ is a combination of veno-arterial compliance ratio (=0.96) and venous compartment resistance (=SVR × 0.038), with the dimensions of resistance, and is based on a formula including age, height, and weight [43]:

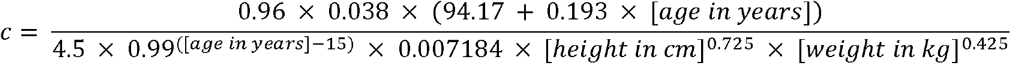

Driving pressure for venous return (VRdP) will be defined as the pressure difference between Pmca and CVP (VRdP = Pmca – CVP). Resistance to venous return (RVR) will be defined as the resistance downstream of Pmca to reflect resistance for venous return and will be calculated as the ratio of the pressure difference between Pmca and CVP and CO [RVR = (Pmca – CVP) / CO]. This formula will be used to describe venous return during transient states of imbalances (Pmca is the average pressure in the circulation and RVR is the resistance encountered to the heart) [44,45].

#### 3.4.3 Efficiency measures

Efficiency of the heart (E_h_) will be defined as the ratio of the pressure difference between Pmca and CVP and Pmca [E_h_ = (Pmca – CVP) / Pmca] (0 ≤ E_h_ ≤ 1). This equation is proposed for the measurement of heart performance, i.e., how well the heart handles the VRdP in terms of Pmca and CVP [39,40,42]. A value ∼1 reflects a normal heart function with CVP close to 0. During the cardiac stop ejection, right atrial pressure (i.e., CVP) is equal to the Pmca, and E_h_ approaches zero [39].

Cardiac power [Power = CO × (MAP – CVP) × 0.0022] and power output [CPO = (CO × MAP) / 451] will be also calculated. Cardiac power represents the rate of energy input the systemic vasculature receives from the heart at the level of the aortic root to maintain the perfusion of the vital organs in shock states [46]. Power efficiency (E_power_) will be defined as the ratio between the change in power and the change in Pmca [E_power_ = Δ((MAP-CVP) × CO) × 0.0022 / ΔPmca]. Whereas E_h_ is a static variable, E_power_ dynamically describes the change in cardiac power in relation to the change in power (MAP × CO) and Pmca [47].

Volume efficiency (E_vol_) will be calculated as the ratio of the pressure difference between Pmca and right atrial pressure (i.e., CVP) and the change in Pmca [E_vol_ = Δ(Pmca-CVP) / ΔPmca] (0 ≤ E_vol_ ≤ 1). Volume efficiency conveys a dynamic variable embodying the efficiency of added fluid, vasopressor, or inotrope in terms of increase in VRdP related to increase in Pmca, and therefore CO and oxygen delivery [47].

#### 3.4.4 Other circuit parameters

We will also calculate arterial compliance [C_art_ = SV / (SAP – DAP)], arterial resistance [R_art_ = MAP / (SV × HR)], venous compartment resistance [R_ven_ = SVR × 0.038], and effective arterial elastance (E_a_ = MAP / SV) which is an integrative measure of cardiac afterload that includes steady and pulsatile components.

### 3.5 Outcomes

#### 3.5.1 Primary outcomes

##### Rationale

Because the study will assess how vasopressor choice influences both cerebral autoregulation and advanced systemic hemodynamics, as well as the interaction between these two domains, the primary outcome comprises two co-primary measures along with an interaction metric.

###### a. Cerebral autoregulation will be assessed by

i. *Intracranial pressure monitoring*, which serves as a surrogate in conjunction with CPP changes. A high ICP that remains relatively stable when CPP fluctuates suggests preserved autoregulation, whereas an ICP that changes proportionally to CPP indicates impaired autoregulation.
ii. *Near-infrared spectroscopy (NIRS)*, using bilateral forehead sensors to measure regional cerebral oxygen saturation (rSO_2_) in the frontal cortex. Changes in rSO_2_ are linked to shifts in cerebral blood flow and are used to estimate autoregulatory function.

From these modalities, the following autoregulation indices will be calculated:

- Pressure reactivity index: a moving Pearson correlation coefficient between 10-s averaged ICP and MAP signals computed over 5-min windows (standard method). Pressure reactivity index reflects cerebrovascular pressure reactivity and is often considered a measure of static autoregulation. A positive PRx indicates impaired autoregulation, whereas a near-zero or negative PRx suggests preserved autoregulation.
- Cerebral oximetry index (COx): a continuous correlation coefficient between spontaneous fluctuations in MAP and rSO□. A stable autoregulatory state results in a near-zero or negative COx value, as rSO□ shows no correlation or a negative correlation with arterial pressure.

###### b. Endpoint metric

The primary cerebral autoregulation endpoint is the area under the PRx versus time curve (AUC) from randomization to 48 hours (lower AUC = better average autoregulation). Mean PRx and the percentage of time PRx ≤ 0 (preserved autoregulation) during this period will also be reported. Cerebral oximetry index will be analyzed in secondary and interaction models to complement PRx-based findings.

###### c. Measurement

continuous ICP and NIRS monitoring; PRx and COx calculated continuously and stored; hourly summary values archived.

###### a. Definitions

i. **Cardiac output** (measured by advanced monitoring device).
ii. **Effective arterial elastance** as an index of net arterial load (calculated from arterial waveform/CO data; Ea = MAP / stroke volume).

###### b. Endpoint metric

Co-primary systemic hemodynamic metric is the combined AUC of CO and Ea over 48 hours (report each separately and a pre-specified composite — e.g., time-weighted average CO and Ea).

###### c. Measurement

continuous/beat-to-beat arterial waveform and CO monitor; hourly summary values archived.

###### a. Definition

the primary interaction analysis tests whether treatment arm modifies the relationship between PRx/COx and CI/Ea — formally: test the treatment × CO (or Ea) interaction term in a mixed-effects model of PRx over time.

###### b. Endpoint metric

significance and effect size of the interaction term over 0-48 hours (i.e., does the relationship between CO/Ea and PRx/COx differ across Group N, V, and NV).

#### 3.5.2 Key secondary outcomes

Secondary outcomes complement the primary focus and provide mechanisms, safety, and clinical context.

##### 1. Extended cerebral physiology

○ **Near-infrared spectroscopy regional oxygen saturation**: continuously measured bilaterally over the frontal cortex. Data will include mean and minimum values, as well as the percentage of time with rSO_2_ below 55%. The COx will be derived to assess autoregulation, reporting both the AUC and the percentage of time with COx values indicating impaired autoregulation. Near-infrared spectroscopy data will be analyzed in conjunction with systemic hemodynamics to evaluate the effects of vasopressors and to explore interactions between autoregulation and hemodynamic variables.

##### 2. Advanced systemic hemodynamics and vascular metrics

○ **Systemic vascular resistance** and **stroke volume variation**: hourly summaries and AUCs 0-48 hrs.
○ **Mean arterial pressure target attainment:** % time within prespecified MAP target window (e.g., 80-100 mmHg).
○ **Vasopressor dose exposure:** cumulative dose (μg kg^-1^) over 48 hours and time-weighted average dose.
○ **Mean arterial pressure variability indices:** e.g., average real variability (ARV) or SD of MAP over intervals.

##### 3. Intracranial and clinical safety

○ **Intracranial pressure:** mean, peak, and time > 20/25 mmHg; ICP AUC 0-48 hrs.
○ **Cerebral perfusion pressure:** mean and time < 60 mmHg.
○ **Adverse events:** arrhythmias requiring treatment, ischemic events (limb/mesenteric), hyponatremia (for AVP), myocardial ischemia, device-related complications.
○ **Organ perfusion markers:** serum lactate, urine output, acute kidney injury (KDIGO criteria).

#### 3.5.3 Short-term clinical outcomes

○ **ICU length of stay**.
○ **Ventilator-free days at 28 days**.
○ **In-hospital mortality**.
○ **Neurological outcome at 3 months** (GOSE) —exploratory if followup feasible.

#### 3.5.4 Exploratory outcomes

○ **Time to restoration of stable autoregulation** (PRx consistently ≤0 for X consecutive hours).
○ **Subgroup analyses**: baseline autoregulation status (preserved versus impaired), age strata, mass lesion versus diffuse injury.

### 3.6 Adherence to protocol

Multiple strategies will be implemented to support recruitment and ensure adherence to the study protocol. Before recruitment begins, and periodically throughout the recruitment phase, trained research coordinators and investigators will actively promote the REGULATE trial and provide education to staff at departmental meetings. Key elements of the study —including eligibility criteria and the details of the interventions— will be reviewed with senior medical personnel in both the ED and ICU. This will help secure support for the trial and ensure that senior clinicians maintain sufficient clinical equipoise to allow eligible patients to be randomized. Core aspects of the inclusion and exclusion criteria, as well as the three interventions, will be repeatedly emphasized to clinical staff to facilitate recruitment and protocol compliance. Randomization packs will be readily available in the ED and ICU and will be regularly restocked by research coordinators. Upon patient randomization, research coordinators will contact the treating clinicians to review and discuss the assigned intervention.

### 3.7 Sample size and predefined statistical analysis plan

The primary outcomes of the study —cerebral autoregulation and advanced systemic hemodynamics— will be analyzed using linear mixed-effects models. Treatment arm, baseline values, and time will be included as fixed effects, and a random intercept for each patient will account for repeated measures. Interaction analyses will assess whether the relationship between PRx and systemic hemodynamic indices (CO or Ea) differs by treatment group by including treatment × CO (or Ea) interaction terms in the models. To account for multiple primary outcomes, a Bonferroni-adjusted significance threshold of α = 0.025 will be applied.

Secondary outcomes, including ICP, CPP, MAP, heart rate, vasopressor exposure, and adverse events, will be analyzed using mixed-effects models for continuous variables and generalized linear models for binary outcomes, with repeated measures and patient-level random effects as appropriate. For secondary endpoints, a significance threshold of α = 0.05 will be used, with the false discovery rate applied to adjust for multiple comparisons.

Missing data will be addressed using multiple imputations under a missing-at-random assumption, with sensitivity analyses conducted using complete-case and alternative imputation approaches. Subgroup analyses will be performed for baseline autoregulation status, TBI severity, age, and relevant comorbidities, including treatment × subgroup interaction terms in the models to evaluate differential treatment effects.

Sample size calculations were based on detecting a 0.05-unit difference in PRx AUC between groups with 80% power, assuming a standard deviation of 0.1 and accounting for multiplicity; a total of approximately 450 patients randomized 1:1:1 across norepinephrine, vasopressin, and combination arms is planned to ensure adequate power for both primary outcomes.

### 3.8 Data collection

Clinical data will be obtained through review of electronic medical records and medical charts. Data analysis will be based on predefined and contemporaneously recorded measurements. The staff will be blinded to measurements until the end of the study and all data are analyzed.

The general characteristics of the patients will be recorded, including demographic information, diagnosis, disease severity (e.g., SOFA and SAPS II scores, where higher scores within 24 hours of enrollment indicate more severe organ dysfunction and higher mortality), APACHE II score (with higher scores reflecting more severe disease, poorer prognosis, and increased mortality), and tissue perfusion parameters.

#### 3.8.1 Measurement timing and data handling

- *Baseline window*: -1 to 0 hour before randomization (when available) — obtain baseline hemodynamic and other parameters.
- *Primary analysis windows*: 0-48 hours (primary), with secondary analyses 0- 48 hours and overall ICU stay.
- *Sampling cadence*: continuous monitoring for PRx/MAP/ICP/other; store raw data or minute/hourly aggregates.
- Quality control: requirement of minimum contiguous monitoring (e.g., ≥36 of 48 hours of acceptable data) for inclusion in primary AUC analyses (for artifact rejection rules see section “3.9 Data monitoring, safety, and management”).

#### 3.8.2 Pre-specified thresholds / definitions

- **Autoregulation failure:** PRx > +0.3 (common threshold); sensitivity analyses with PRx > +0.2 and > +0.4 will be also reported.
- **Hemodynamic instability:** MAP < 65 mmHg and/or CO < 4.0 L min^-1^ for > 2 minutes.

### 3.9 Data monitoring, safety, and management

An independent Data Safety Monitoring Committee (DSMC), comprising experts in critical care medicine, biostatistics, and clinical trials, will oversee the safety, ethical, and scientific aspects of the study. The DSMC was convened during the protocol development phase and will conduct one interim analysis after day-30 follow-up is completed for the first 50 patients to monitor recruitment, protocol adherence, and safety. Based on this review, the DSMC will advise whether the trial should continue as planned.

All monitoring devices will be calibrated according to manufacturer instructions and will not display treatment labels, while vasopressor syringes will be prepared by an unblinded pharmacist who will not participate in the study. If the attending physician at any time deems that any vasopressor is ineffective, switching types of vasopressors or adding a second vasopressor will be allowed. Patients who will be switched from norepinephrine to AVP and vice versa will be defined as ‘‘refractory’’ to that agent.

The goal of the clinical data management plan is to ensure high-quality data by implementing standardized procedures that minimize errors and missing information, thereby generating an accurate database for analysis. Remote monitoring will be employed to detect early aberrant patterns, inconsistencies, credibility issues, and other anomalies.

#### 3.9.1 Artifact rejection rules

To ensure the validity of cerebral autoregulation and systemic hemodynamic measurements, predefined artifact rejection rules will be applied. Data will first be screened for physiological plausibility, excluding values outside accepted ranges (ICP < 0 or > 60 mmHg, MAP < 40 or > 160 mmHg, CPP < 30 or > 120 mmHg, CO < 2.00020or > 12.0 L min^-1^, rSO_2_ < 20% or > 95%). Signals demonstrating abrupt changes unlikely to be physiologic, including a ≥40 mmHg change in SAP, DAP, or MAP within two minutes, or ICP/MAP fluctuations >30% within 30 seconds, will be discarded. Segments showing loss of pulsatility, disconnection, damping artifacts, or device-flagged poor signal quality (e.g., NIRS flat line or sensor off) will also be excluded.

For autoregulation indices, PRx will be calculated as a moving Pearson correlation coefficient between 10-second averaged ICP and MAP values over 5-minute windows; any window with >20% rejected data will be excluded from analysis. Similarly, COx will be derived from MAP and rSO_2_ signals, with exclusion of windows containing >20% low-quality or missing NIRS data. At the patient level, a minimum of 36 out of 48 hours of acceptable data will be required for inclusion in primary analyses. Within hourly epochs, at least 80% valid data will be required for retention.

### 3.10 Ethics and dissemination

The study will be approved by the Ethics Committee of Tzaneio General Hospital and conducted in accordance with the Declaration of Helsinki, the International Conference on Harmonization guideline for Good Clinical Practice, and applicable local regulations. The trial will be registered on ClinicalTrials.gov. Written informed consent will be obtained from each patient or their legally authorized representative before initiation of any study procedure. The final study dataset will be available to investigators and stored for 5 years after trial completion. Study results will be submitted to peer-reviewed journals and presented at scientific conferences, with publication expected in 2026-2027.

#### 4. Screening and enrollment workflow

##### i. Patient arrival

○ Patient with severe TBI presents to ED or ICU.

##### ii. Initial assessment for eligibility (ED or ICU)

○ Confirm GCS ≤ 8 after resuscitation.
○ Confirm age ≥ 18 years.
○ Assess need for vasopressor therapy to maintain CPP/MAP.
○ Evaluate contraindications (see exclusion criteria).

##### iii. Screening documentation (ED or ICU)

○ Clinical staff record inclusion/exclusion criteria in screening log.
○ Assign screening ID for tracking.

##### iv. Informed consent (ED or ICU)

○ Obtain consent from legally authorized representative.

##### v. Randomization 1:1:1 (ICU)

○ Randomization via computer-generated permuted block list with concealed allocation.

- Eligible and consented patients randomized to:
- Group N: Norepinephrine infusion 0.1–0.5 μg kg^-1^ min^-1^
- Group V: AVP infusion 0.01–0.06 U min^-1^
- Group NV: Combination norepinephrine and AVP

##### vi. Baseline assessment (ICU)

○ Record demographics, injury characteristics, baseline ICP, MAP, PRx, CO, Ea, and other monitoring data.
○ Verify feasibility of continuous ICP and arterial line monitoring.

##### vii. Blinding (ICU)

○ Quadruple blinding maintained: patients/families, evaluators, data analysts, and follow-up staff.

##### viii. Intervention and monitoring (ICU)

○ Continuous ICP, MAP, CO, Ea, and other hemodynamic monitoring initiated.
○ Vasopressor dose adjusted per protocol to maintain target CPP/MAP.
○ PRx and advanced hemodynamic indices recorded continuously for primary endpoint analysis (0-48 hours).

##### ix. Follow-up and data collection (ICU, post-ICU)

○ Secondary outcomes collected over 0-48 hours.
○ Clinical outcomes (GOSE, ICU length of stay, mortality) collected per protocol.

##### x. Analysis (ICU, post-ICU)

○ Intention-to-treat principle applied.
○ Data included in mixed-effects models and other pre-specified statistical analyses.

## 5. Discussion

Traumatic brain injury remains a leading cause of morbidity and mortality worldwide, with impaired cerebral autoregulation recognized as a major determinant of secondary brain injury and poor long-term outcomes. Despite advances in multimodal monitoring, there is limited high-quality evidence to guide the choice of vasopressor therapy for optimizing cerebral and systemic hemodynamics in this patient population. The REGULATE trial has been designed to address this knowledge gap by directly comparing norepinephrine, AVP, and their combination with respect to cerebral autoregulation and advanced systemic cardiovascular performance in patients with severe TBI.

The pathophysiological rationale for REGULATE is strong. Norepinephrine is the most widely used vasopressor in neurocritical care; however, experimental and clinical data suggest that its effects on cerebral autoregulation are dose-dependent and may be detrimental in vulnerable patients. Vasopressin, with distinct receptor selectivity and systemic hemodynamic effects, may represent a mechanistically favorable alternative, yet its cerebral vascular impact is poorly characterized. By incorporating continuous monitoring of autoregulation indices (PRx and COx), NIRS, and advanced systemic hemodynamic measurements, REGULATE will provide a comprehensive evaluation of how these agents influence both cerebrovascular reactivity and cardiovascular coupling in the acute phase of severe TBI.

The study design has several strengths. First, the use of a randomized controlled, quadruple-blinded methodology minimizes bias and ensures robust causal inference. Second, the co-primary endpoints —autoregulation and systemic hemodynamic metrics— reflect the dual physiological domains most relevant to secondary injury prevention. Third, predefined interaction analyses will clarify how systemic hemodynamic states modify the relationship between vasopressor therapy and autoregulatory function, advancing the field beyond descriptive associations. Finally, the planned large sample size provides adequate statistical power to detect clinically meaningful differences and allows for rigorous subgroup analyses.

There are, however, potential limitations. Given the open-label nature of vasopressor administration to treating clinicians, treatment titration may be influenced by clinical judgment, though outcome assessors, analysts, and follow-up staff remain blinded. The complexity of multimodal monitoring may introduce technical variability and data loss, although rigorous artifact rejection protocols and quality control measures have been pre-specified. Furthermore, while the study is powered for physiological endpoints, clinical outcomes such as mortality and neurological recovery are secondary, and definitive conclusions regarding long-term benefit will require subsequent confirmatory trials.

In summary, REGULATE is the first adequately powered trial to systematically evaluate the comparative effects of norepinephrine, AVP, and their combination on cerebral autoregulation and systemic hemodynamics in severe TBI. By integrating advanced monitoring and comprehensive outcome assessment, this study is expected to generate pivotal mechanistic insights and inform evidence-based vasopressor strategies in neurocritical care. Ultimately, the results have the potential to guide individualized hemodynamic management and reduce the burden of secondary brain injury in this highly vulnerable population.

## Data availability

Data will be made available upon request after publication through a collaborative process. Researchers should provide a methodically sound proposal with specific objectives in an approval proposal.

## Acknowledgements

Nothing to acknowledge.

## Author contributions

AC contributed to study design, protocol draft, review and editing of the manuscript.

## References

1. Aries MJ, Elting JW, De Keyser J, Kremer BP, Vroomen PC. Cerebral auto-regulation in stroke: a review of transcranial Doppler studies. Stroke. 2010;41(11):2697–704.

2. Brady KM, Lee JK, Kibler KK, Easley RB, Koehler RC, Shaffner DH. The lower limit of cerebral blood flow autoregulation measured by indocyanine green kinetics during hypo- and hypercarbia. Anesth Analg. 2008;106(1):327– 33.

3. Steiner LA, Czosnyka M, Piechnik SK, Smielewski P, Chatfield DA, Menon DK, et al. Continuous monitoring of cerebrovascular pressure reactivity allows determination of optimal cerebral perfusion pressure in head-injured patients. Crit Care Med. 2002;30(4):733–8.

4. Jaeger M, Schuhmann MU, Soehle M, Meixensberger J. Clinical significance of impaired cerebrovascular autoregulation after severe brain trauma. J Neurol Neurosurg Psychiatry. 2006;77(4):515–6.

5. Czosnyka M, Smielewski P, Kirkpatrick P, Menon DK, Pickard JD. Monitoring of cerebral autoregulation in head-injured patients. Stroke. 1996;27(10):1829–34.

6. Zeiler FA, Ercole A, Cabeleira M, Menon DK, Smielewski P, Czosnyka M. Cerebral autoregulation in clinical practice: a review of the literature. Anesth Analg. 2019;128(5):1181–94.

7. Froese L, Dian J, Gomez A, Unger B, Zeiler FA. The cerebrovascular response to norepinephrine: A scoping systematic review of the animal and human literature. Pharmacol Res Perspect. 2020 Oct;8(5):e00655. doi: 10.1002/prp2.655. PMID: 32965778; PMCID: PMC7510331.

8. Vedrenne-Cloquet M, Chareyre J, Léger PL, Genuini M, Renolleau S, Oualha M. Low Dosing Norepinephrine Effects on Cerebral Oxygenation and Perfusion During Pediatric Shock. Front Pediatr. 2022 Jul 6;10:898444.

9. Meng L, Sun Y, Zhao X, Rasmussen M, Al-Tarshan Y, Meng DM, Liu Z, Adams DC, McDonagh DL. Noradrenaline-induced changes in cerebral blood flow in health, traumatic brain injury and critical illness: a systematic review with meta-analysis. Anaesthesia. 2024 Sep;79(9):978–991.

10. Sedhai YR, Shrestha DB, Budhathoki P, Memon W, Acharya R, Gaire S, Pokharel N, Maharjan S, Jasaraj R, Sodhi A, Kadariya D, Asija A, Kashiouris MG. Vasopressin versus norepinephrine as the first-line vasopressor in septic shock: A systematic review and meta-analysis. J Clin Transl Res. 2022 May 25;8(3):185–199.

11. Heming N, Mazeraud A, Azabou E, Moine P, Annane D. Vasopressor Therapy and the Brain: Dark Side of the Moon. Front Med (Lausanne). 2020 Jan 10;6:317.

12. Moppett IK, Sherman RW, Wild MJ, Latter JA, Mahajan RP. Effects of norepinephrine and glyceryl trinitrate on cerebral haemodynamics: transcranial Doppler study in healthy volunteers. Br J Anaesth. 2008 Feb;100(2):240–4.

13. Salvagno M, Geraldini F, Coppalini G, Robba C, Gouvea Bogossian E, Annoni F, Vitali E, Sterchele ED, Balestra C, Taccone FS. The Impact of Inotropes and Vasopressors on Cerebral Oxygenation in Patients with Traumatic Brain Injury and Subarachnoid Hemorrhage: A Narrative Review. Brain Sci. 2024 Jan 24;14(2):117.

14. Permpikul C, Tongyoo S, Viarasilpa T, Trainarongsakul T, Chakorn T, Udompanturak S. Early Use of Norepinephrine in Septic Shock Resuscitation (CENSER). A Randomized Trial. Am J Respir Crit Care Med. 2019 May 1;199(9):1097–1105.

15. Huh J, Kwon H, Park H, Park SC, Yun SS, Chae MS. Impact of Norepinephrine and Dopamine Infusion on Renal Arterial Resistive Index during Pre-Emptive Living Donor Kidney Transplantation: Propensity Score Matching Analysis. Medicina (Kaunas). 2024 Jun 28;60(7):1066.

16. Stocchetti N, Maas AI, Chieregato A, van der Plas AA. Hyperventilation in head injury: a review. Chest. 2005 May;127(5):1812–27.

17. Babar SI, Berg RA, Hilwig RW, Kern KB, Ewy GA. Vasopressin versus epinephrine during cardiopulmonary resuscitation: a randomized swine outcome study. Resuscitation. 1999 Jul;41(2):185–92.

18. Jozwiak M. Alternatives to norepinephrine in septic shock: Which agents and when? J Intensive Med. 2022 Jun 12;2(4):223–232.

19. Walum H, Westberg L, Henningsson S, Neiderhiser JM, Reiss D, Igl W, Ganiban JM, Spotts EL, Pedersen NL, Eriksson E, Lichtenstein P. Genetic variation in the vasopressin receptor 1a gene (AVPR1A) associates with pairbonding behavior in humans. Proc Natl Acad Sci U S A. 2008 Sep 16;105(37):14153–6.

20. Valverde A, Giguère S, Sanchez LC, Shih A, Ryan C. Effects of dobutamine, norepinephrine, and vasopressin on cardiovascular function in anesthetized neonatal foals with induced hypotension. Am J Vet Res. 2006 Oct;67(10):1730–7.

21. Fage N, Asfar P, Radermacher P, Demiselle J. Norepinephrine and Vasopressin in Hemorrhagic Shock: A Focus on Renal Hemodynamics. Int J Mol Sci. 2023 Feb 17;24(4):4103.

22. Froese L, Dian J, Batson C, Gomez A, Alarifi N, Unger B, Zeiler FA. The Impact of Vasopressor and Sedative Agents on Cerebrovascular Reactivity and Compensatory Reserve in Traumatic Brain Injury: An Exploratory Analysis. Neurotrauma Rep. 2020 Nov 6;1(1):157–168.

23. Highton D, Ghosh A, Tighe D, Uff C, Al-Rawi P. Dynamic cerebral autoregulation after traumatic brain injury. Br J Anaesth. 2020;125(2):200–8

24. Silver JM, McAllister TW, Yodofsky SC, eds. Textbook of Traumatic Brain Injury. Arlington, Va: American Psychiatric Publishing; 2005, pp. 27–39.

25. Teasdale G, Jennett B. Assessment of coma and impaired consciousness: a practical scale. Lancet 1974;2:81–4.

26. Meyfroidt G, Bouzat P, Casaer MP, Chesnut R, Hamada SR, Helbok R, Hutchinson P, Maas AIR, Manley G, Menon DK, Newcombe VFJ, Oddo M, Robba C, Shutter L, Smith M, Steyerberg EW, Stocchetti N, Taccone FS, Wilson L, Zanier ER, Citerio G. Management of moderate to severe traumatic brain injury: an update for the intensivist. Intensive Care Med. 2022 Jun;48(6):649–666.

27. Wiles MD. Management of traumatic brain injury: a narrative review of current evidence. Anaesthesia. 2022 Jan;77 Suppl 1:102–112.

28. Lavinio A, Coles JP, Robba C, Aries M, Bouzat P, Chean D, Frisvold S, Galarza L, Helbok R, Hermanides J, van der Jagt M, Menon DK, Meyfroidt G, Payen JF, Poole D, Rasulo F, Rhodes J, Sidlow E, Steiner LA, Taccone FS, Takala R. Targeted temperature control following traumatic brain injury: ESICM/NACCS best practice consensus recommendations. Crit Care. 2024 May 20;28(1):170.

29. Slot RER, Helbok R, van der Jagt M. Update on traumatic brain injury in the ICU. Curr Opin Anaesthesiol. 2025 Apr 1;38(2):93–99.

30. Robba C, McCredie V, Chesnut RM, Citerio G, Gauss T, Hawryluk GWJ, Helbok R, Meyfroidt G, Newcombe V, Sarwal A, Taccone FS, van der Jagt M, Wahlster S, Zanier ER, Bouzat P. Traumatic brain injury management in the intensive care unit: standard of care and knowledge gaps. Intensive Care Med. 2025 Jun;51(6):1112–1127.

31. Bögli SY, Olakorede I, Beqiri E, Chen X, Lavinio A, Hutchinson P, Smielewski P. Cerebral perfusion pressure targets after traumatic brain injury: a reappraisal. Crit Care. 2025 May 21;29(1):207.

32. Tsigaras ZA, Weeden M, McNamara R, Jeffcote T, Udy AA; PRECISION-TBI Investigators. The pressure reactivity index as a measure of cerebral autoregulation and its application in traumatic brain injury management. Crit Care Resusc. 2023 Dec 14;25(4):229–236.

33. Huijben JA, Dixit A, Stocchetti N, Maas AIR, Lingsma HF, van der Jagt M, et al.; CENTER-TBI Investigators and Participants. Use and impact of high intensity treatments in patients with traumatic brain injury across Europe: a CENTER-TBI analysis. Crit Care 2021;25:78.

34. Ferreira FL, Bota DP, Bross A, Melot C, Vincent JL. Serial evaluation of the SOFA score to predict outcome in critically ill patients. JAMA 2001;286:1754–8.

35. Perel P, Edwards P, Shakur H, Roberts I. Use of the Oxford Handicap Scale at hospital discharge to predict Glasgow Outcome Scale at 6 months in patients with traumatic brain injury. BMC Med Res Methodol 2008;8:72.

36. Gardner RM. Direct blood pressure measurement–dynamic response requirements. Anesthesiology 1981;54:227–36.

37. Saugel B, Kouz K, Scheeren TWL, et al. Cardiac output estimation using pulse wave analysis-physiology, algorithms, and technologies: a narrative review. Br J Anaesth 2021;126:67–76.

38. Hamzaoui O, Monnet X, Richard C, et al. Effects of changes in vascular tone on the agreement between pulse contour and transpulmonary thermodilution cardiac output measurements within an up to 6-hour calibration-free period. Crit Care Med 2008;36:434–40.

39. Chalkias A, Laou E, Mermiri M, et al. Microcirculation-guided treatment improves tissue perfusion and hemodynamic coherence in surgical patients with septic shock. Eur J Trauma Emerg Surg 2022;48:4699–711.

40. Chalkias A, Laou E, Papagiannakis N, et al. Assessment of Dynamic Changes in Stressed Volume and Venous Return during Hyperdynamic Septic Shock. J Pers Med 2022;12:724.

41. Parkin WG, Leaning MS. Therapeutic control of the circulation. J Clin Monit Comput 2008;22:391–400.

42. Chalkias A, Laou E, Papagiannakis N, et al. Determinants of venous return in steady-state physiology and asphyxia-induced circulatory shock and arrest: an experimental study. Intensive Care Med Exp 2022;10:13.

43. Wijnberge M, Sindhunata DP, Pinsky MR, et al. Estimating mean circulatory filling pressure in clinical practice: a systematic review comparing three bedside methods in the critically ill. Ann Intensive Care 2018;8:73.

44. Berger D, Moller PW, Takala J. Reply to “Letter to the editor: Why persist in the fallacy that mean systemic pressure drives venous return?”. Am J Physiol Heart Circ Physiol 2016;311:H1336–7.

45. Berger D, Moller PW, Weber A, et al. Effect of PEEP, blood volume, and inspiratory hold maneuvers on venous return. Am J Physiol Heart Circ Physiol 2016;311:H794–806.

46. Fincke R, Hochman JS, Lowe AM, et al; SHOCK Investigators. Cardiac power is the strongest hemodynamic correlate of mortality in cardiogenic shock: a report from the SHOCK trial registry. J Am Coll Cardiol 2004;44:340–8.

47. Sondergaard S, Larsson JS, Möller PW. The haemodynamic effects of crystalloid and colloid volume resuscitation on primary, derived and efficiency variables in post-CABG patients. Intensive Care Med Exp 2019;7:13.

48. Tejerina EE, Pelosi P, Robba C, Penuelas O, Muriel A, Barrios D, et al; VENTILA Group. Evolution over time of ventilatory management and outcome of patients with neurologic disease. Crit Care Med 2021;49:1095–106.

49. Holland MC, Mackersie RC, Morabito D, Campbell AR, Kivett VA, Patel R, et al. The development of acute lung injury is associated with worse neurologic outcome in patients with severe traumatic brain injury. J Trauma 2003;55:106–11.

50. Mascia L, Sakr Y, Pasero D, Payen D, Reinhart K, Vincent JL; Sepsis Occurrence in Acutely Ill Patients (SOAP) Investigators. Extracranial complications in patients with acute brain injury: a post-hoc analysis of the SOAP study. Intensive Care Med 2008;34:720–7.

51. Contant CF, Valadka AB, Gopinath SP, Hannay HJ, Robertson CS. Adult respiratory distress syndrome: a complication of induced hypertension after severe head injury. J Neurosurg 2001;95:560–8.

52. Mascia L, Zavala E, Bosma K, Pasero D, Decaroli D, Andrews P, et al.; Brain IT Group. High tidal volume is associated with the development of acute lung injury after severe brain injury: an international observational study. Crit Care Med 2007;35:1815–20.

53. Mascia L, Fanelli V, Mistretta A, Filippini M, Zanin M, Berardino M, et al. Lung-Protective Mechanical Ventilation in Patients with Severe Acute Brain Injury: A Multicenter Randomized Clinical Trial (PROLABI). Am J Respir Crit Care Med 2024;210:1123–31.

54. Wang L, Zhou XH, Richardson TS. Identification and estimation of causal effects with outcomes truncated by death. Biometrika 2017;104:597–612.

